# The association between suicidal ideation with future suicide attempts and service utilization: a population cohort study in Ontario, Canada

**DOI:** 10.1101/2024.12.25.24319633

**Authors:** C. Chan, W. Wodchis, P. Kurdyak, P. Donnelly

## Abstract

**Background:** Few studies examine suicidal ideation and its relationship with suicide attempts and health service use using a community sample.

**Objective:** To examine the association between suicidal ideation with (i) future mental health service use, and (ii) future suicide attempts, in a general population sample.

**Methods:** Using the 2002 and 2012 Canadian Community Health Surveys – Mental Health cycles linked with health administrative data, this study followed 14,708 Ontarians 15 years and older in a retrospective cohort study to examine patterns of mental health service use within 1 year, and suicide attempts within 5 years of self-reported suicidal ideation.

**Results:** A total of 2.1% (n=302) of all survey respondents reported suicidal ideation alone in the past year and another 0.5% (n=76) reported having ideation-with-action (i.e. ideation with planning or previous suicide attempt). Ideation-with-action was associated with a higher likelihood of future suicide attempts and future mental health service use, when compared to suicidal ideation alone. However, the majority of survey respondents who attempted suicide within the five-year follow-up period did not report a previous attempt or ideation at baseline.

**Conclusion:** While suicidal ideation is statistically significantly associated with greater likelihood of suicide attempt within five years, most attempters did not self-report ideation at baseline. Further study is needed to measure suicidal outcomes longitudinally to allow for dynamic expression of suicidal ideation as outcomes such as suicide attempts are measured.

Those with ideation-with-action are more intense users of mental health services than those with ideation alone, indicating that care is reaching the more severe ideators.

## Introduction

Suicidal ideation is the third most important predictor of suicide after mental health hospitalizations and prior suicide attempts (Franklin et al, 2017; World Health Organization, 2014). Only a small proportion of those with ideation will act on their thoughts (Maris, 1981) so there is a need to understand which types of people with ideation are most likely to act, and in what circumstances. Among ideators, the conditional probability of transitioning to making a plan is about one third (34%) and of ever making an attempt has been reported to be 29% (Nock et al, 2008a).

Current suicide theories distinguish suicidal ideation from suicidal behaviour and employ ‘ideation-to-action models’ to emphasize that risk factors for ideation differ from risk factors for suicide attempt (Klonsky et al, 2018). However, studies differentiating those with ideation from those who have attempted or will attempt suicide are few (May & Klonsky, 2016), and where studies exist, they tend to focus on subpopulations such as veterans (Veterans Affairs Canada, 2019) or students (Liu et al, 2023) and do not provide general community-based estimates. There is a need to assess the value of using ideation as a predictive screen for suicide risk in a general community population.

### The role of mental health services

Suicidal ideation is related to psychological distress and disorder or unmanageable emotional distress (Dour et al, 2011; Shneidman, 1996) and affected individuals could benefit from mental health care treatment. Therefore, understanding potential associations between ideation and service utilization can help assess whether use of mental health resources when responding to suicidal ideation and attempts is impactful on subsequent progression to action.

Evidence for the effectiveness of mental-health services in lowering suicidality in the general population is, however, unclear. A scoping review on the link between mental health service availability and suicide rates among community-dwelling populations resulted in heterogeneous findings (Chan et al, 2024). There was a near even split on the number of positive (36%) and negative (34%) correlations, with almost one third (30%) generating null correlations: meaning that some studies found that greater availability of mental health services were associated with lower suicide rates (a desirable correlation), some studies found greater availability of mental health services were associated with higher suicide rate (a correlation counter to expectation), and some studies could not confirm any correlation. While the positive associations between mental health services and suicide may be suggestive of reverse causation where perhaps policies and services were appropriately introduced to match high rates of suicide in a jurisdiction, the large number of null findings renders it impossible to make any conclusions at a system level, also due to differences in study designs, most of which were cross-sectional in nature.

Alongside availability of care determined by jurisdictions, predisposing, enabling and need factors also influence an ideator’s help-seeking behaviour, as per the Andersen Model of Health Care Utilization (Andersen, 1995).

Better understanding of the link between suicidal ideation and attempts must also explore the ideation-to-action models by considering ideation alone versus ideation accompanied by action, whether this be suicide planning or attempt. This approach adds to knowledge on which specific subgroups of ideators would most benefit from treatment.

### Research objective

This study explored the risks associated with suicidal ideation in terms of future mental health service use among community-dwelling individuals who report suicidal ideation, and future suicide attempts.

## Methods

### Study population and data sources

The study population was created by pooling Ontario respondents from two cross-sectional surveys: the 2002 and 2012 Canadian Community Health Surveys – Mental Health (CCHS-MH) administered by Statistics Canada between May and December 2002, and between January and December 2012. The surveys used a multistage sample allocation strategy to gather data from a randomly selected representative sample of households across Canada and included modules on health status, mental well-being, mental illness, determinants of mental health, demographics and other variables (Statistics Canada, 2014; Beland, 2002). Participants were aged 15 years and older living in private dwellings in Canada’s ten provinces covering approximately 98% of Canada’s population. Detailed methodology is available from Statistics Canada reports (2004, 2013a, 2013b, 2014).

The overall survey response rate was 77% for the 2002 survey (CCHS, 2002) and 69% for the 2012 survey (CCHS, 2013).

Statistics Canada sought consent from CCHS participants to share their survey responses with provincial ministries of health and link responses to administrative databases. All Ontario CCHS-MH respondents from 2002 and 2012 cycles who consented to linkage of their data to health administrative databases available through Ontario’s single-payer healthcare system were eligible for this study, enabling longitudinal measurement of health-care service utilization and mortality patterns over time at ICES. In total, 79% consented to database linkage. Multiple health administrative databases, linked together using unique encoded identifiers, were used for this study.

The Canadian Community Health Surveys (Mental Health component) were linked to multiple administrative databases using unique encoded identifiers and analysed at Institute for Clinical Evaluative Sciences (ICES) in Toronto, Ontario. ICES is an independent, non-profit research institute in Ontario. It uses population-based health information to produce knowledge on a wide range of healthcare issues.

Ontario’s Registered Persons Database (RPDB) contains sociodemographic information on persons registered under the Ontario Health Insurance Plan (OHIP). Physician visit data was obtained from the OHIP database; emergency department visit information was obtained from the Canadian Institute for Health Information (CIHI) National Ambulatory Care Reporting System (NACRS), and hospitalization data were obtained from the CIHI Discharge Abstract Database (DAD) and the Ontario Mental Health Reporting System (OMHRS). See Appendix 3 for details on databases used in this study.

### Cohort creation

Using all Ontario CCHS-MH respondents from 2002 and 2012 survey cycles, participants successfully linked to the RPDB were included, and after removing those with missing data for the exposure variable, 14,708 were included in the analysis (**Figure 4.1**).

**Figure 4.1:**
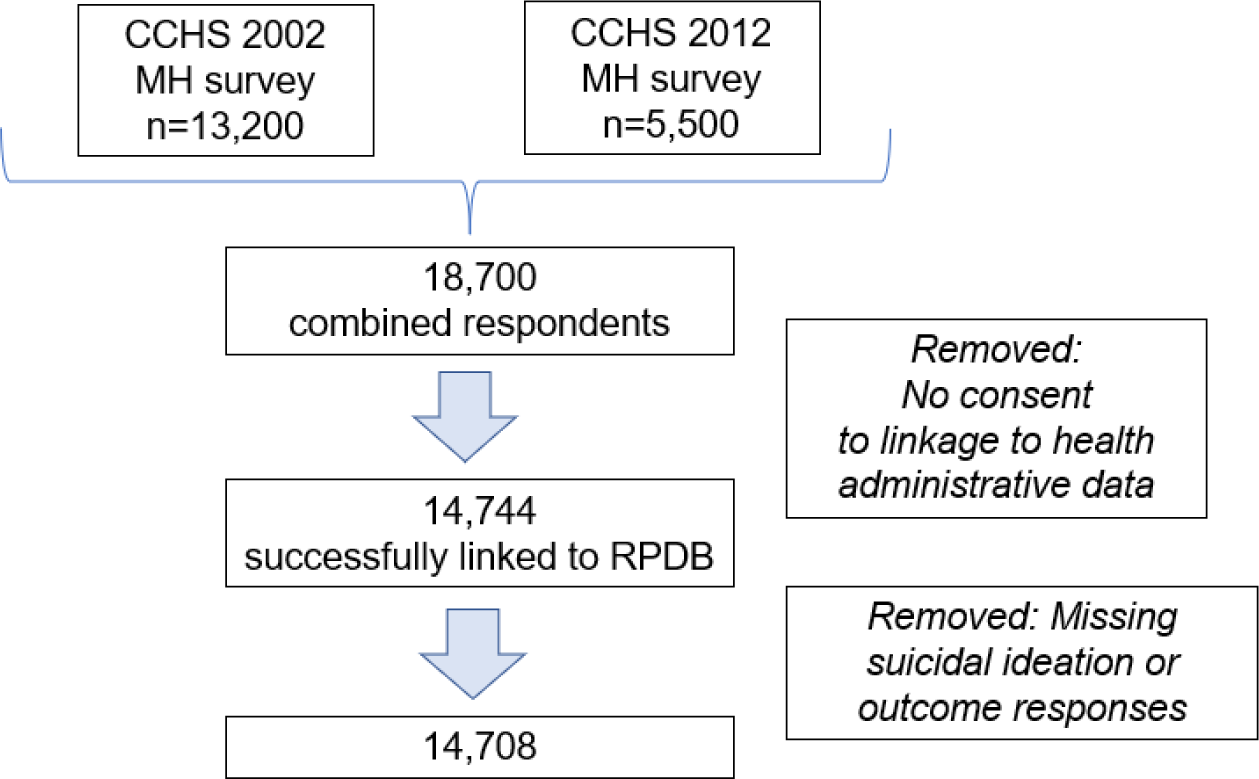
Flow chart of study population from pooled Ontario component of Statistics Canada’s Canadian Community Health Surveys (CCHS) Mental Health component, linked to health administrative data through the Registered Persons Database (RPDB).

### Exposure

The exposure variable, suicidal ideation, was created by identifying persons with: (i) no ideation; (ii) ideation alone; and (iii) ideation with a suicidal plan or with a previous suicide attempt.

In the CCHS, respondents were asked about suicidal ideation only if they answered yes to a subclinical depression question “Did you feel sad, empty or depressed most of the day, nearly every day, during (a) period of 2 weeks?”.

*i. Suicidal ideation (past 12 months)*: All CCHS respondents were prompted with the question about lifetime suicide ideation “Have you ever seriously thought about committing suicide or taking your own life?” and if they answered yes, then they were asked *“Has this happened in the past 12 months?”* with two response options: *Yes, No.* Respondents were classified as reporting ideation if they answered “yes” to the latter question.
*ii. Suicidal ideation with plan*: All CCHS respondents who answered affirmatively to ideation in the past 12 months were prompted with the question *“In the past 12 months”, “you made a plan for committing suicide?”* with two response options: *Yes, No.* Respondents were classified as having ideation with a plan if they responded “yes”.
*iii. Suicidal ideation with past-year suicide attempt:* All CCHS respondents who answered affirmatively to ideation in the past 12 months were prompted with the question *“During the past 12 months”, “you attempted suicide or tried to take your own life?”* with two response options: *Yes, No.* Respondents were classified as having ideation with a past-year attempt if they responded “yes”.

For brevity, respondents who answered yes to ii and iii above are considered to have ‘ideation-with-action’ in this study.

### Outcomes

The study population was followed 1 year from CCHS-MH interview date for health utilization and 5 years from CCHS-MH interview date for suicide attempts. Outcome measures were defined as follows.

*Outpatient visits:* **Mental health-related outpatient visits** consisted of outpatient visits to a psychiatrist for any reason or non-psychiatrist (i.e. primary care outpatient visits to family physician) for any mental health condition, within 1 year following CCHS interview date. These are based on both diagnostic and counselling codes.

*Acute-care visits:* **Mental health-related acute care visits** consisted of **emergency department** visits (coded with a primary diagnosis of *International Statistical Classification of Diseases and Related Health Problems, 10th Revision (ICD-10)* F06-F99, X60-X84, Y10-Y19, or Y60-X84, and Y28 as a secondary diagnosis with no F06-F99 as a primary diagnosis) within 1 year following CCHS interview date, or **hospitalizations** defined as (1) inpatient admissions with primary diagnosis of ICD-10 F06-F99 or any diagnosis of X60-X84, Y10-Y19, and Y28 as a secondary diagnosis with no F06-F99 as a primary diagnosis; or (2) an admission in the Ontario Mental Health Reporting System; within 1 year following CCHS interview date.

An emergency department (ED) visit that led to a hospital admission was considered a single hospitalization; to avoid double-counting, ED visits reflect visits without hospital admission.

*Suicide attempts:* **Suicide attempt-related visits** consisted of emergency department visits for deliberate self-harm; ICD-10 codes X60-84 (intentional self-harm), Y10-19 (poisoning of undetermined intent), Y28 (contact with sharp object, of undetermined intent), Y87 (late effects of intentional self-harm) including visits with suspect diagnoses (suspect=T), within 5 years following CCHS interview date. Studies have endorsed the use of emergency department data for identifying deliberate self-harm (Bethell and Rhodes, 2009).

With regard to the follow-up periods, health service utilization was measured within one year of the baseline suicidal ideation measure in keeping with existing literature (Gilbody et al, 2002; Ghanbari et al, 2015). The study intended to measure suicide attempts in the same observation period but due to the small number of events, it was necessary to follow respondents for five years following index survey date.

Note also that there were too few completed suicides to examine in this cohort.

### Covariates

Sociodemographic covariates were **age, sex, marital status, income, education and employment status** as reported on the CCHS survey. These variables were chosen as they have been shown to be important risk factors for both suicidal ideation and suicide (**Appendix 1**).

Two health-related covariates were analysed:

### i. Mood and anxiety disorders

Mood and anxiety disorders were defined in the CCHS by using the Diagnostic and Statistical Manual of Mental Disorders, IV Edition (DSM-IV) criteria (American Psychiatric Association, 2000) and were assessed by using a Canadian adaptation of the World Mental Health Composite International Diagnostic Interview (WMH-CIDI), which is a structured interview for psychiatric disorders (Statistics Canada, 2013a; Kessler and Ustun, 2004). Diagnostic algorithms identified respondents meeting the criteria for the measured mood and anxiety disorders (major depressive episode, bipolar I, bipolar 2, hypomania, or phobia, obsessive-compulsive disorder, agoraphobia, panic disorder, social phobia) in the past 12 months. For example, a diagnosis of major depressive episode requires at least one episode of two weeks or more of persistent low mood, loss of interest or pleasure in normal activities with associated changes in sleep pattern, changes in appetite, feelings of guilt, hopelessness, impaired concentration, or suicidal thoughts (Statistics Canada, 2013b).

### ii. Substance use disorders

Substance use disorder includes alcohol and drug use disorders and was measured in the CCHS using the modified WMH-CIDI algorithms derived from DSM-IV applied to symptom data to define substance use disorders, that is, substance abuse and/or dependence for alcohol, cannabis and other drugs, in the past 12 months. Substances covered were cannabis, cocaine, amphetamines, MDMA (ecstasy), hallucinogens, solvents, heroin, and steroids.

**Self-reported help-seeking** was also studied as a covariate. The CCHS collects information about respondent’s use of help, and health care services related to mental health problems (i.e. problems with emotions, mental health, or use of alcohol or drugs) during the past 12 months through this question “Have you ever seen, or talked on the telephone, to any of the following professionals about your emotions, mental health or use of alcohol or drugs?” which was asked of all respondents. The professionals included psychiatrists, family doctors or general practitioners, other medical doctors, psychologists, nurses, social workers, counsellors or psychotherapists, religious or spiritual advisors, and other professionals. “Other professionals” referred to acupuncturists, biofeedback teachers, chiropractors, energy healing specialists, exercise or movement therapists, herbalists, homeopaths or naturopaths, hypnotists, guided imagery specialists, massage therapists, relaxation experts, yoga or meditation experts, and dieticians.

### Statistical analysis

The distributions of mental healthcare service use and self-harm outcomes were examined along with the covariates, stratified by suicidal ideation exposure (no ideation, ideation, and ideation-with-action i.e. with plan or attempt). Baseline characteristics of the exposure groups were compared using unweighted frequencies and weighted percentages for categorical variables. Chi-square tests were used to compare difference in proportions between the subgroups since all covariates were categorical variables. Statistical significance was indicated by a 2-tailed *P* value < 0.05. When the *P* value was significant, a post-hoc test was performed to find out which cells from the contingency table were different from their expected values, to identify which cell made the largest contribution to the result (Beasley et al, 1995).

Factors significantly associated with suicidal ideation in univariate analyses (p < 0.05) were included in multivariable models as covariates/confounders. Multiple logistic regression was conducted to identify the predictive characteristics of ideation-with-action compared to ideation alone (for mental health service use and suicide attempts); this was important in order to study the difference between ideators and actors of suicidality.

Odds and rate ratios of having a subsequent mental health visit and of having a subsequent self-harm episode were estimated using health administrative data, and corresponding 95% Wald confidence intervals (CI) were generated using multiple logistic regression and negative binomial regression models. These are shown in **Tables 4.2** and **4.3**. The Negative Binomial model was used for the outpatient visits outcome due to overdispersion and a large number of zeros or no encounters in the data (Hilbe, 2007). The possibility of multicollinearity between the variables of interest was examined using the variance inflation factor (VIF) indicator; there was no indication of multicollinearity as the VIF of all variables of interest was <5 (Kim, 2019). Correlation matrices were also examined, and elimination and combination of variables tested, to assess for collinearity.

Joint effects of ideation and mental health service utilization were of interest in this study but the low number of events rendered it impossible to conduct a study whereby subgroups of similar risk profiles would be followed up to examine which individuals attempt suicide, taking into account varying levels of health service utilization. Therefore, models were run to examine, separately, the likelihood of health service utilization and of suicide attempt.

Sampling weights provided by Statistics Canada were applied in all analyses to account for the multistage survey sampling design and to ensure that estimates were representative of the Ontario population.

Sensitivity analyses were used to examine differences between the two survey waves, and the impact of excluding observations with missing data (see **Appendix 2**.)

All analyses were performed with SAS version 9.4 (SAS Institute Inc, Cary, North Carolina).

### Ethics

The survey datasets used in this study were accessed at ICES, an independent, non-profit research institute in Ontario which uses population-based health information to produce knowledge on a wide range of healthcare issues. The study cohort consists of CCHS respondents who consented to have their survey data shared with ICES for linkage with health administrative data. The use of these data for health system planning and evaluation was authorized under Section 45 of Ontario’s Personal Health Information Protection Act, which allows for administrative approval and does not require review by a research ethics board.

Data for the study was accessed for research purposes from 26 April 2023 to 26 September 2024. Authors did not have access to information that could identify individual participants during or after data collection.

## Results

### Univariate analyses

A total of 2.1% of community-dwelling survey respondents (n= 302) reported suicidal ideation alone in the past year and another 0.5% reported having ideation-with-action i.e. with a plan or attempt (n=76).

As shown in **Table 4.1**, subgroups did not differ in terms of sex, education, and employment. However, there were statistically significant differences between the groups in other characteristics: compared to persons with ideation alone, those with ideation-with-action tended to be younger, poorer, unpartnered, have a substance use disorder, and mood and anxiety disorder (p < 0.01).

**Table 4.1:**
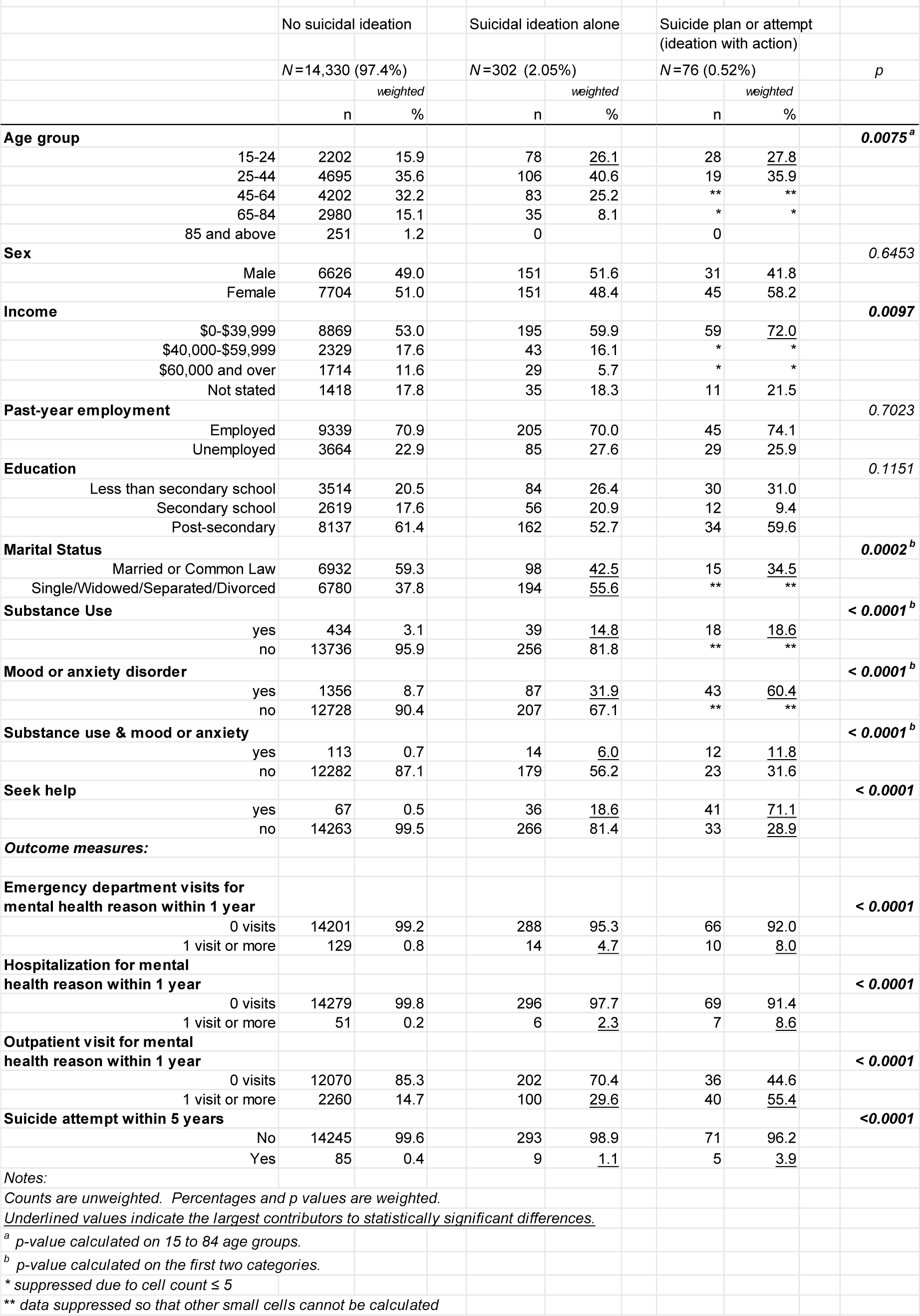
Baseline characteristics of those without ideation, those with ideation alone, and those with suicide plan or attempt.

The prevalence of reporting having both mood and anxiety disorder and substance use disorder was twice as high in the ideation-with-action subgroup (12%) compared to the ideation-alone subgroup (6%).

Compared to persons with ideation alone, those with ideation-with-action used more health services including outpatient visits, hospitalization and ED visits within a year of reporting suicidal ideation (p < 0.0001).

Compared to persons with ideation alone, those with ideation-with-action had a higher rate of suicide attempts within 5 years of self-reporting ideation (p < 0.0001).

However, most (86%) of those who attempted suicide were non-ideators at baseline (n=85).

So while the larger number of suicide attempts were amongst non-ideators, the proportion of this group who attempted suicide was much lower than among ideators.

The results indicate that only 19% of persons with ideation alone self-reported seeking help for mental health problems compared to a greater proportion (71%) in the ideation-with-action subgroup.

### Multivariable analyses

Multiple logistic regression was conducted to study the characteristics of persons with ideation-with-action, compared to persons with ideation alone. This is to address the gap in literature in understanding how risk factors for ideation alone could be different from risk factors for suicide attempts (Klonsky et al, 2018). Although ideation alone carries its own mental burden which warrants support and treatment, ideation-with-action (i.e. ideation with planning or a previous suicide attempt) increases risk of future suicide attempts and deaths (Franklin et al, 2017).

From the logistic regression model (**Table 4.2**), all variables were statistically significantly different when comparing individuals with ideation alone to individuals with ideation-with-action.

**Table 4.2:** Associations between sociodemographic factors, health conditions, and help-seeking, with suicidal ideation with planning or previous attempt (ideation with action) compared to suicidal ideation alone.

|  |  | Ideation alone (n=302) vs Ideation-with-action (n=76) |  |  |  |
| --- | --- | --- | --- | --- | --- |
|  |  | Crude |  | Adjusted |  |
|  |  | OR | (95%, CI) | OR | (95%, CI) |
| **Sociodemographic factors** |  |  |  |  |  |
| **Age group** |  |  |  |  |  |
|  | 15-24 | 1.3* | (1.3, 1.4) | 1.5* | (1.4, 1.5) |
|  | 25-44 | 1.3* | (1.3, 1.4) | 0.9* | (0.9, 0.9) |
|  | 45-84 (Ref.) |  |  |  |  |
| **Sex** |  |  |  |  |  |
|  | Male | 0.9* | (0.9, 0.9) | 1.1* | (1.0, 1.1) |
|  | Female (Ref.) |  |  |  |  |
| **Income** |  |  |  |  |  |
| | \$0-\$39,999 | 2.6* | (2.5, 2.6) | 1.2* | (1.2, 1.3) |
| | \$40,000-\$59,999 | 1.3* | (1.2, 1.3) | 1.1 | (1.1, 1.2) |
| | \$60,000 and over (Ref.) | | | | |
| **Past-year employment** |  |  |  |  |  |
|  | Yes | 0.8* | (0.8, 0.8) | 0.7* | (0.7, 0.8) |
|  | No (Ref.) |  |  |  |  |
| **Education** |  |  |  |  |  |
|  | Less than secondary school | 1.2* | (1.1, 1.2) | 1.6* | (1.5, 1.6) |
|  | Secondary school | 0.5* | (0.4, 0.5) | 0.7* | (0.7, 0.8) |
|  | Post-secondary (Ref.) |  |  |  |  |
| **Marital Status** |  |  |  |  |  |
|  | Married/Common Law | 0.5* | (0.4, 0.5) | 0.5* | (0.5, 0.6) |
|  | Single/Widowed/Divorced (Ref.) |  |  |  |  |
| **Health factors** |  |  |  |  |  |
| **Substance Use** |  |  |  |  |  |
|  | Yes | 1.4* | (1.4, 1.4) | 1.3* | (1.3, 1.4) |
|  | No (Ref.) |  |  |  |  |
| **Mood and anxiety disorder** |  |  |  |  |  |
|  | Yes | 5.0* | (4.9, 5.1) | 2.3* | (2.2, 2.4) |
|  | No (Ref.) |  |  |  |  |
| **Help-seeking factors** |  |  |  |  |  |
| **Seek help** |  |  |  |  |  |
|  | Yes | 11.8* | (11.5, 12.1) | 10.1* | (9.8, 10.4) |
|  | No (Ref.) |  |  |  |  |
| *p < 0.05 |  |  |  |  |  |

Mood and anxiety disorder and marital status yielded the highest adjusted odds ratios. Overall, marital status (aOR 0.5 [0.5, 0.6]) and having a job (aOR 0.7 [0.7, 0.8]) were protective factors, while the adjusted odds for mood and anxiety disorder (aOR 2.3 [2.2, 2.4]), younger age (aOR 1.5 [1.4, 1.5]), less education (aOR 1.6 [1.5, 1.6]), substance use disorder (aOR 1.3 [1.3, 1.4]), and lower income (aOR 1.2 [1.2, 1.3]) were much higher among those with ideation-with-action, compared to those with ideation alone.

The adjusted odds of those with ideation-with-action self-reporting that they sought help for mental health problems was ten times those with ideation alone (aOR 10.1 [9.8, 10.4]) suggesting that those with more severe ideation are appropriately seeking and accessing care.

Multivariable analyses were conducted to examine actual mental health service use and suicide attempts in the two ideation subgroups (ideation-with-action versus ideation alone) controlling for confounders identified as being statistically significant from univariate analyses. Results are shown in **Table 4.3**.

**Table 4.3:** Association of mental health service utilization, and of suicide attempt, with those with suicidal ideation with planning or previous attempt (ideation with action) compared to ideation alone.

|  |  |  |  |  |
| --- | --- | --- | --- | --- |
|  | Ideation alone (n=302) vs Ideation-with-action (n=76) |  |  |  |
|  | Crude OR | (95%, CI) | Adjusted OR | (95%, CI) |
| Emergency department mental health visit (1 or more events v no events) | 1.7* | (1.7, 1.8) | 1.2* | (1.2, 1.3) |
| Mental health hospitalization (1 or more events v no events) | 3.9* | (3.8, 4.1) | 3.3* | (3.1, 3.5) |
| Outpatient mental health visit (1 or more events v no events) | 1.6** | (1.6, 1.6) | 1.5** | (1.5, 1.5) |
| Suicide attempt after 5 years | 1.8* | (1.7, 2.0) | 1.2* | (1.1, 1.3) |
| *p < 0.05 using logistic regression. |  |  |  |  |
| **p < 0.05 using negative binomial regression. |  |  |  |  |
| All adjusted results controlled for age, income, marital status, substance use, mood and anxiety disorder, and self-reported help-seeking. |  |  |  |  |

With regard to service utilization, hospitalization yielded the highest adjusted odds ratio. Those with ideation-with-action were more likely to have been hospitalized (aOR 3.3 [3.1, 3.5]) and to have emergency-department visits (aOR 1.2 [1.2, 1.3]) within one year of follow-up, compared to those with ideation alone. The rate of outpatient visits for those with plan or attempt was 50% higher than for those with ideation alone (IRR = 1.5 [1.5, 1.5], p <0.0001).

With regard to suicide attempt, the odds for those with ideation-with-action to attempt suicide within 5 years were 1.2 times (aOR 1.2 [ 1.1, 1.3]) higher than those with ideation alone.

## Discussion

One in 50 CCHS respondents had suicidal ideation (2.1%), and one in 200 (0.5%) had a plan or previous suicide attempt. The main variables associated with having a plan or past attempt are male, being unpartnered, younger age, unemployed, less educated, no job, less income, having a mood or anxiety disorder, a substance use disorder and help-seeking for mental health problems. The results also show that there are many more individuals with no suicidal ideation who attempt suicide in the subsequent five years, suggesting that while the presence of suicidal ideation might be associated with subsequent suicide attempts, the absence of ideation does not preclude the later development of suicidality. Put another way, suicidal ideation is a state, not a trait (Joiner, 2005).

### Prevalence of suicidal ideation

The prevalence rate of 2.1% past-year ideation in this study equates to about a million Canadians considering ending their lives each year and is in line with the WHO World Mental Health Survey which noted a 12-month prevalence of ideation at 2% and suicide attempts at 0.3% in high-income countries (Borges et al, 2010).

### Sociodemographic factors

Multivariable analysis of this Ontario population found that younger persons had higher adjusted odds of ideation with planning or previous attempt, compared to older persons. This finding is consistent with research with global trends showing that younger age confers higher suicide risk (Sinyor et al, 2017). An American general-population study found that past-year suicidal ideation, planning and attempt was higher among younger (18-39 years old) than older (≥40 years old) persons (Ivey-Stephenson et al, 2022) but contrasts with a study of nine countries and more than 50,000 participants which found that the 12-month suicidal ideation rate is significantly higher for older people than for younger people but there is no difference in the 12-month suicide attempt rate for older and younger people (Cabello et al, 2020).

Being partnered was a key protective factor and was strongly associated with lower odds of ideation-with-action, compared to ideation alone – a finding which concurs with other studies which highlight loss of connectedness and support including marital status as risk factors for suicidal ideation and behaviours (Liu et al, 2022; van Wijingaarden et al, 2014; King and Merchant, 2008; Pompili et al, 2007) in line with the Interpersonal Theory of Suicide (Joiner, 2005) and the Integrated Motivational-Volitional (IMV) model (O’Connor, 2011).

### Health conditions

The odds of self-reporting mood-and-anxiety disorders were 2.3 times (and for self-reporting substance use disorder, 1.3 times) among those with ideation-with-action compared to having ideation alone. These findings concur with studies that found depression to be the most critical risk factor for attempting suicide (Miller et al, 2020; Yang et al, 2015; Oquendo et al, 2004; Thomas and Morris, 2003) and that suicide risk increases with substance use (Swann et al, 2022; Ashrafioun et al, 2017; Wilcox et al, 2004).

### Self-reported help-seeking for mental health problems

The finding that most of the future attempters (92%) did not self-report seeking professional help for mental health problems in the CCHS is consistent with Canadian studies which found that only a small proportion of people with suicidal ideation or who have attempted suicide seek help (Cheung and Dewa, 2007; Farand et al, 2004). Self-reported help-seeking for mental health problems was ten times more likely among those with ideation-with-action compared to ideation alone, suggesting that higher-suicide-risk individuals are making good choices to reach out for help.

These patterns are likely reflective of the predisposing, enabling and need elements of the Andersen Model of Health Care Utilization (Andersen, 1995). For example, those with ideation alone may not contemplate seeking help due to low levels of mental-health or suicide literacy (Gabriel and Violato, 2010) and misunderstanding of depressive symptoms and suicidality (Oliffe et al, 2016). In contrast, those with ideation-with-action may seek help in an effort to prevent acting on their thoughts, perhaps from knowing the impact from their previous suicidal actions and having experienced the morbidity related to ideation that is beyond just thinking. Indeed, a Canadian study found that attempters have a higher perception of need than those with ideation only (Pagura et al, 2009).

It is also possible that those with lower severity of ideation may be appropriately assessing informal mental health support such as family, friends, school, workplace, or church support which is not measured in the ‘help-seeking’ variable focused on professional help.

Apart from individual-level factors, important external factors such as the responsible or irresponsible reporting of media in reporting suicides can also affect help-seeking (Niederkrotenthaler et al, 2023) and social media content (Sinyor et al, 2023) but were not measured in this study.

### Mental health service utilization from health administrative data

This study found that after controlling for covariates, suicidal ideation-with-action did predict actual mental health utilization, a finding replicated in other studies (Pirkis et al, 2001a). Specifically, suicidal ideation-with-action was associated with greater likelihood of using mental health services than ideation alone, in all settings.

The study also found that 30% of ideators had at least one outpatient visit in the year after self-reported ideation, and 55% of planners/attempters had at least outpatient visit in the year after self-reported ideation. These findings suggest a substantial proportion of ideators do not receive outpatient care for mental health reasons. Even if these individuals did not attempt suicide, it is likely an indication of under-treatment as these individuals are likely suffering lower health status and quality of life associated with ideation.

For planners/attempters, the effect of increased and actual use of all mental health services (compared to those with ideation alone) remained significant after controlling for a range of potential confounders including: sociodemographic factors (age, marital status and income), substance use and mood/anxiety disorders. This suggests that ideators who are not clinically depressed, but who experience sadness, emptiness or subclinical depression (pre-requisites for the suicidal ideation question in the survey), and who have either a plan or previous attempt, have higher use of mental health services. Thus, ideation can also be a general marker of emotional and psychological distress driving an individual to seek help (Nock et al, 2008b; Maris, 1981).

Despite this finding, the study found low rates of mental health service use across the board: when looking at ideators-without-action, the majority did not use mental health services (95%, no ED visits; 98% no hospitalizations; and 70% no outpatient visits, for mental health problems), and among ideators-with-action, large proportions did not use mental health services (92% no ED visits; 91% no hospitalizations; and 45% no outpatient visits). This is of concern considering 47% of ideators-without-action, and 79% of ideators-with-action, self-reported having either a mood or anxiety or substance use disorder.

### Suicide attempts

This study found an association between suicidal ideation and suicide attempts. Suicide attempts among those with ideation-with-action were 1.2 times more likely compared to those with ideation alone. While 20% higher odds are not large, the direction of the finding is consistent with evidence in the literature that previous suicide attempt is a predictor of future suicide attempts (Bostwick et al, 2016). Other studies have shown that those with past-year suicidal ideation have significantly higher past-year prevalence rates of suicide attempts at 15% in high-income countries and that suicidal planning further increases this risk (Nock et al, 2013; Borges et al, 2010); and that previous history of attempts is one of the most robust predictors of future attempts and suicide (Fedyszyn et al, 2016; Ribeiro et al, 2015; Hawton et al, 2015).

### Most attempters do not have a previous attempt history

This section, along with the next two sections on “attempters” refers to those persons who attempted suicide as identified by health administrative data i.e. individuals presenting to hospital.

The majority of attempters (86%) did not have any previous history of attempts, which is in line with a review finding that most attempters do so once in their life: 23% re-attempt non-fatally but 70% make no further attempt (Owens, 2002).

### A large proportion of attempters have no mental disorders

Among attempters, 41% did not self-report having a mental disorder (neither a mood or anxiety disorder nor a substance use disorder) reinforcing the findings of previous studies suggesting sizable proportions of attempters do not have mental disorders (Oquendo et al, 2024) and emphasizing that attention needs to be paid to attempters who do not have a mental disorder and, with that, the role of subclinical depression (Joiner et al, 2017).

Additionally, this study found that among individuals with ideation, those with ideation-with-action tend to have a co-occurring mood and anxiety and substance use disorders which presents an opportunity for health professionals to explore suicide risk during medical appointments, increase mental health/suicide literacy, promote positive views of services and encourage seeking support for suicidal ideation.

### Attempters mostly do not receive mental health services

Most attempters (92%) did not self-report help-seeking for mental health problems – despite more than half self-reporting having a mental disorder – is provocative. In terms of actual mental health use within 1 year of ideation-disclosure, most (77%) did not have any hospitalizations, most (62%) did not have any ED visits and a large proportion (42%) did not have any outpatient visits for mental health reasons. This also means 58% did access outpatient services within a year of follow-up despite most (92%) self-reporting no help-seeking for mental health problems in the previous year.

This is consistent with a US general population study which found that less than half of attempters had clinical contact around the time of their attempt (Bommersbach et al, 2022b).The authors emphasized that while suicide attempts have increased in the US, efforts need to be paid to increase service use among high-risk individuals, especially as suicide attempt is a very strong risk factor for suicide (Favril et al, 2022).

### Strengths of the study

The follow-up design of a randomly selected and representative community sample is the most notable and distinctive strength of this study.

The large provincially representative study links multiple cycles of the Canadian Community Health Survey with health administrative databases to evaluate the association between suicidal ideation, service utilization and suicidality outcomes. It was advantageous to be able to pool two surveys together to increase the size of the study population to study rare outcomes and to take advantage of the dataset linkages to explore baseline self-reported characteristics alongside health-system visits. Additionally, use of a large sample allowed for sub-group analyses with sufficient statistical power.

Insights into participants’ sociodemographic characteristics and history of mental health conditions, albeit self-reported, allowed for more meaningful analyses in that we were able to control for the presence of substance use and mood and anxiety disorders, as well as sociodemographic confounders. Historically, follow-up studies in suicide have often used psychiatric populations which limits generalization to the whole population.

Few studies have examined ideators according to whether there was any planning or suicidal behaviour compared to ideation alone, as was done in this study. For example, some studies combine suicidal ideation and suicide attempts in one group despite these two groups have differential associations with service use and future suicidal behaviour.

### Limitations of this study

This study has a number of important limitations.

Results are not directly applicable to subpopulations that were excluded from the CCHS sampling frame; unrepresented populations include indigenous populations living on reserve, individuals in the military, and those living in institutions. This exclusion is important since it is known, for example, that indigenous persons experience ideation differently (Bolton et al, 2014; Simpson et al, 2003) and indigenous Canadians have three times the suicide rate of non-indigenous Canadians (Kumar and Tjepkema, 2019).

Only individuals who satisfied subclinical depression probe questions were asked about ideation, on the grounds that those who had not been sad, empty or subclinically depressed in the preceding 12 months would have been unlikely to consider suicide. The cohort therefore excludes persons who may have suicidal ideation despite not meeting this threshold. This conditional questioning results in an underestimation of the true prevalence of ideation in the general population.

There are important measurement limitations, particularly that identification of individuals reporting ideation, planning and attempts relied on self-report, with no external validation.

The study examined past-year status of persons who self-reported having disorders covered in the CCHS survey. Persons who do not meet the criteria for the disorders covered by the survey may still have another type of mental disorder, and others may not meet the criterion of past-year and could have had disorders earlier in life.

The study measured baseline suicidal ideation, a measure of suicide risk at a single point in time, and categorizes respondents into ‘no ideators’, ‘ideators only’ and ‘ideators-with-action’. Treating these categories as fixed may result in underestimating suicide risk in some since suicidality is a phenomenon that is transient and fluctuating (Bornheimer et al, 2022). The study did not measure suicidal ideation over time; therefore, whether individuals with no baseline suicidal ideation subsequently developed suicidal ideation cannot be established with certainty, including among those who had a suicide attempt in the 5-year follow-up period. That individuals did not report suicidal ideation at baseline but had a subsequent suicide attempt during the follow-up period likely means they developed suicidal ideation during the follow-up period and they may or may not have sought help for this.

Mental-health related visits with types of care providers which are not captured by the administrative databases could not be accounted for during follow-up, which could confound any reported associations. Finally, there was no measurement of the quality of care provided to those with ideation, which was beyond the scope of the study and data sources to assess. Thus, care may be suboptimal.

There are also analytical limitations. Due to splitting of the study population into small samples according to ideation subgroups, some of the final samples did not have an adequate statistical power to detect effect sizes of small or moderate magnitude.

Despite the large datasets used, there were too few suicides from the data sources to examine suicides in this study. Similarly, there were too few suicide attempts within 1 year and 3 years of follow-up and sufficient numbers only at the 5-year point, yet the small number of events even at the 5-year point limits our ability to make inferences about the relationship between ideation and suicide attempts.

The study design necessarily excludes the ability to measure subtle or time-dependent factors that impact suicidal behaviour such as grief, recent loss, stressors, financial woes, which were not measured in the data sources.

### Conclusion

This study found that most of the future suicide attempters from the CCHS did not admit to ideation at baseline but it also found that ideation-with-action is more strongly related to future attempt, compared to ideation alone. There is therefore a risk between baseline suicidal ideation and attempt, and given the long (5-year) follow-up period, it is unsurprising that there were attempters among those with no baseline suicidal ideation even though the prevalence is low relative to the total sample.

The clinical utility of suicidal ideation for prediction of suicide attempts among those with baseline suicidal ideation has merit but the large number of individuals who attempted suicide with no baseline suicidal ideation suggests that the clinical utility is limited to those with measurable suicidal ideation, and not those without. Further study is needed to measure suicidal outcomes longitudinally to allow for dynamic expression of suicidal ideation as outcomes such as suicide attempts are measured.

The most intense use of mental health services was among individuals with ideation-with-action whether or not they had a mental health disorder, suggesting that ideators who might not be clinically depressed but who experienced sadness, emptiness and subclinical depression, have increased access to services. Compared to those with ideation alone, persons with ideation-with-action tend to have a co-occurring mood and anxiety and substance use disorders which presents an opportunity for health professionals to explore suicide risk during medical appointments and encourage help-seeking.

## Data Availability

The data are held at Institute of Clinical Evaluative Sciences in Ontario, Canada.

## Appendices

### Appendix 1: Literature justifying the selection of covariates

This study used the following **sociodemographic covariates**: age, sex, marital status, income, education and employment status. Suicide rates have been shown to differ by sex and age: men account for three times the number of suicides than women; suicide rates are highest in adults aged 70 and older across both men and women, and a disproportionately large number take place in younger age groups such as 15 to 29 (WHO, 2014; Patton et al, 2009; Nock et al, 2008a). With regard to ideation specifically (rather than suicide deaths), there is research showing that females had greater levels of suicidal ideation using validated scales, compared to men (Park et al, 2005) but also mixed findings on whether ideation was more prevalent among females or males (Ibrahim et al, 2017; Lewinsohn et al, 2001).

In general, suicide studies do not tend to compare younger to older age groups and instead focus on one age group, highlighting the need for future research since at-risk age groups might differ, especially for cross-country comparisons (Nock et al, 2009). Our study includes logistic regression analyses comparing younger with older age groups and adds to this gap in the literature.

With regard to marital status, literature suggests that separation, divorce and widowhood may increase the risk of suicide (Kolves et al, 2010; Yeh et al, 2008; Brockington, 2001).

With regard to income, research shows that income is inversely associated with psychological distress (McMillan et al, 2010; Lynch et al, 2000; Ferrada-Noli, 1997).

With regard to education, education has been shown to be related to suicidality through the life course in that those with lower levels of education have the highest suicide rates (Phillips and Hempstead, 2017; Lorant et al, 2005).

With regard to employment, a systematic review found that unemployment is associated with greater incidence of suicide (Milner et al, 2014; Milner et al, 2013).

**Mood and anxiety disorders** were investigated as a covariate because of the strength of evidence in the literature on the impact of mood and anxiety disorders on suicidality and its morbid and mortality outcomes. We know that most people with psychiatric conditions do not die by suicide. However, some psychiatric conditions are more strongly associated with suicidal behaviours than others, for example, major depressive episodes account for at least half of suicide deaths (Holma et al, 2014) and is strongly correlated with ideation, attempt and planning (Melhem et al, 2019; Gill et al, 2018; Large, 2016; Nepon et al, 2010; Bertolote and Fleishmann, 2002).

**Substance use and dependence** was investigated as a covariate also because of the strength of evidence in the literature on the impact of substance abuse on suicidality and its morbid and mortality outcomes. Many studies have shown that suicide risk is elevated in connection to substance use disorders, increasing risk of ideation and behaviour. Suicide death is up between ten to 15 times higher in people with alcohol use disorder and opioid use disorder compared to the general population (Swann et al, 2022; Ashrafioun et al, 2017; Wilcox et al, 2004).

Evidence of the relationship between alcohol use and suicide (HHS, 2012; Pompili et al, 2010) exists but is limited. For example, it is known that acute alcohol intoxication is present in about 30 to 40% of suicide attempts (SAMHSA, 2009; Cherpitel et al, 2004). Evidentiary basis for the relationship between drug misuse and suicide is even less explored (SAMHSA, 2015).

There are studies have shown that substance use when present with other mental disorders escalates suicidal behaviour (Nock et al, 2013; Vijayakumar et al, 2011). It is thought that substance use disorder may interact with depression to increase the risk of engaging in suicidal behaviour (Hoertel et al, 2015). Some studies have shown alcohol consumption, specifically, increases the probability of suicide (Edwards et al, 2022; Kim et al, 2021; Rizk et al, 2021) while a meta-analysis found minimal or negligible differences between these two groups (May & Klonsky, 2016) or found mental disorders in general are minimally or negligibly distinguished these two groups (Kessler et al, 1999). Comorbidity of substance use disorder and mood and anxiety disorder as substance use and major depression has been found to increase the risk of suicide attempts (Bachman et al, 2018; Ginley and Bagge, 2017; Beautrais et al, 1996).

Lastly, self-reported **help-seeking** was included a covariate. The question covered help received in the past 12 months for problems with emotions, mental health or use of alcohol or drugs (referred to in the survey as “mental health problems”) and was asked of all respondents. Although it is possible that help-seeking for suicidal ideation may be similar to help-seeking for other mental health issues, there is little research on how help-seeking behaviour might be different for those with suicidal ideation (Ko, 2018). What we do know is that a large proportion of those with suicidal ideation do not seek help. For example, an American study of adults with suicidal thoughts or past suicide attempts found that fewer than two-thirds received treatment in the past year (Ahmedani et al, 2012). This is consistent with a worldwide study that found that fewer than 20% of those with ideation sought help (Bruffaerts et al, 2011).

### Appendix 2: Sensitivity Analyses

Sensitivity analyses were conducted to determine if important differences exist between the 2002 and 2012 surveys, since the study relies on pooling together respondents from these two surveys. Analyses were also conducted to examine the impact of excluding observations with missing data.

### Numbers of respondents self-reporting past-year suicidal ideation

The 2012 survey included respondents who self-reported suicidal ideation during the worst episode of feeling depressed in the universe from which the question of past-year suicidal ideation was asked. The 2002 survey did not. However, the proportion of respondents that had past-year suicidal ideation from 2002 is 24% and this is comparable to the proportion who had past-year suicidal ideation from 2012 (27%) despite the 2002 survey not including the individuals with suicidal ideation during worst/bad episode into the universe for the past-year suicidal ideation question.

### Numbers of respondents reporting seeking professional help

Among those with ideation, in the 2002 survey, the proportion of respondents who reported seeking professional help - which includes visiting ED, hospitalization, walk-in clinic, consulting a family doctor or psychologist in person or by telephone – was 9% (n=20) and in the 2012 survey, that proportion was 29% (n=47). This three-fold increase in proportion who sought help may indicate cultural trends that facilitate help-seeking in the 2012 sub-sample, such as de-stigmatization of mental problems, and is a factor to be mindful of when considering the pooled population of this study.

### Sociodemographic variables

Ensuring the pooled population from the two surveys are not dissimilar in terms of sociodemographic variables is important since we know, for example, that sex is a factor in suicidal behaviour, with higher rates of ideation and attempts among females (Borges et al, 2010; Nock et al, 2008a). In terms of sociodemographic variables, both cycles had similar proportions of respondents in terms of age, sex, marital, and income. In terms of mood and anxiety disorders, and in substance use disorders, 2002 and 2012 survey respondents had similar proportions.

None of the differences above were of concern in terms of affecting the final results of the study.

### Missing data

Categorizing missing sociodemographic variables (age, sex, marital status, income, education and employment) as separate categories for analysis resulted in no meaningful differences.

Where missing data were few, the cases were deleted from the analytic dataset, with no major impact to the result.

### Appendix 3: Databases used

| Database name | Description |
| --- | --- |
| Canadian Community Health Survey – Mental Health focus | A cross-sectional survey conducted in 2002 and 2012 by Statistics Canada that used a multistage sample allocation strategy to gather data from a randomly selected representative sample of households across Canada and included modules on health status, mental well-being, mental illness, determinants of mental health, health care utilization, demographics and other variables (Statistics Canada, 2014; Beland, 2002). Participants were aged 15 years and older living in private dwellings in Canada’s ten provinces covering approximately 98% of Canada’s population.<br>See: 2012 survey [Surveys and statistical programs - Canadian Community Health Survey - Mental Health and Well-being (CCHS) (statcan.gc.ca)](http://www.statcan.gc.ca/surveys/cchs)<br>2012 survey [Surveys and statistical programs - Canadian Community Health Survey - Mental Health (CCHS) (statcan.gc.ca)](http://www.statcan.gc.ca/surveys/cchs) |
| Registered Persons Database (RPDB) | The RPDB provides basic demographic information (age, sex, location of residence, date of birth, and date of death for deceased individuals) for those issued an Ontario health insurance number. The RPDB also indicates the time periods for which an individual was eligible to receive publicly funded health insurance benefits and the best-known postal code for each registrant on July 1st of each year. See: ICES Data Dictionary for RPDB |
| National Ambulatory Care Reporting System (NACRS) | The NACRS is compiled by the Canadian Institute for Health Information and contains administrative, clinical (diagnoses and procedures), demographic, and administrative information for all patient visits made to hospital- and community-based ambulatory care centres (emergency departments, day surgery units, hemodialysis units, and cancer care clinics). At ICES, NACRS records are linked with other data sources (e.g. DAD, OMHRS) to identify transitions to other care settings, such as inpatient acute care or psychiatric care. See: ICES Data Dictionary for NACRS |
| Discharge Abstract Database (DAD) | The DAD is compiled by the Canadian Institute for Health Information and contains administrative, clinical (diagnoses and procedures/interventions), demographic, and administrative information for all admissions to acute care hospitals, rehab, chronic, and day surgery institutions in Ontario. At ICES, consecutive DAD records are linked together to form “episodes of care” among the |
|  | hospitals to which patients have been transferred after their initial admission. See: ICES Data Dictionary for RPDB |
| Ontario Mental Health Reporting System (OMHRS) | The OMHRS is compiled by the Canadian Institute for Health Information and contains administrative, clinical (diagnoses and procedures), demographic, and administrative information for all admissions to adult designated inpatient mental health beds. This includes beds in general hospitals, provincial psychiatric facilities, and specialty psychiatric facilities. Clinical assessment data is ascertained using the Resident Assessment Instrument for Mental Health (RAI-MH), but different amounts of information are collected using this instrument depending on the length of stay in the mental health bed. Multiple assessments may occur during the length of a mental health admission. See: ICES Data Dictionary for OMHRS |
| Ontario Health Insurance Plan Claims Database (OHIP) | The OHIP claims database contains information on inpatient and outpatient services provided to Ontario residents eligible for the province's publicly funded health insurance system by fee-for-service health care practitioners (primarily physicians) and "shadow billings" for those paid through non-fee-for-service payment plans. The main data elements include patient and physician identifiers (encrypted), code for service provided, date of service, associated diagnosis, and fee paid. See: ICES Data Dictionary for OHIP |

